# Electron microscopy visualization of cell-free mitochondrial DNA–containing extracellular vesicles in human plasma, serum, and saliva

**DOI:** 10.1101/2025.10.15.25338094

**Authors:** Alexandra Volos, Soah Grace Franklin, Jeremy Michelson, Shannon Rausser, Jonathan R. Brestoff, Martin Picard

## Abstract

Human biofluids contain cell-free mitochondrial DNA (cf-mtDNA) and extracellular mitochondria (ex-Mito), creating the challenge of defining their origins, destinations, mechanisms of regulation, and purposes. To expand our understanding of cf-mtDNA biology, we present a descriptive electron microscopy analysis of circulating particles from cf-mtDNA-enriched plasma (citrate, heparin, and EDTA), serum (red and gold top), and saliva collected from ten healthy people (5 females, 5 males, mean age 44.9 years). Ex-mito and extracellular vesicles (EVs) were isolated by centrifugation followed by size-exclusion chromatography, imaged by transmission electron microscopy, and morphometrically analyzed. In parallel, cf-mtDNA was quantified in each biofluid. The resulting catalog of the most common circulating particles in plasma, serum, and saliva show that circulating double-membrane extracellular particles—consistent with mitochondrial ultrastructure—are present across human biofluids, along with EVs and other particle types. Combining imaging with cf-mtDNA quantification, we show that individuals with higher plasma cf-mtDNA concentrations tend to contain more double-membrane, ex-Mito-like particles. These preliminary results challenge the notion that, under normal conditions, the majority of cf-mtDNA exists as naked and potentially pro-inflammatory forms. Instead, these results are consistent with the concept of mitochondria transfer or signaling between cells and tissues. The image inventory provided here expands our knowledge of cell-free mitochondrial biology and provides a resource to inform biofluid selection and technical considerations in future studies quantifying ex-Mito and cf-mtDNA.

## 1. Introduction

Extracellular mitochondria (ex-Mito) function as active mediators of bioenergetic transfer and intercellular signaling, raising the question of which regulated mechanisms facilitate their movement between cells^1^. Understanding these mechanisms requires a systematic framework for categorizing how ex-Mito are transferred. A recent consensus statement^2^ provided such a framework, highlighting that cells do not need direct physical contact to exchange mitochondria. Instead, contact-independent mitochondria transfer can occur when donor cells release ex-Mito into biofluids, which can then be taken up by other cell types within a given tissue or be released into circulation for delivery to cell types in other organs^3,4^.

There are two defined categories of ex-Mito: free mitochondria and extracellular vesicles containing mitochondria (EV-Mito)^2^. Cf-mtDNA can exist either within ex-Mito or as mtDNA fragments released from the mitochondria^3–6^. Fragmented cf-mtDNA is associated with inflammatory conditions^7^; however, cf-mtDNA does not appear to be pro-inflammatory when encased in whole mitochondria^8,9^. These findings have led to the idea that the presence of ex-Mito may indicate the activation of cellular repair processes involving intercellular signaling and mitochondria transfer^10^.

Biofluid selection is a meaningful parameter for cf-mtDNA and ex-Mito research. Platelet activation leads to an increased cf-mtDNA count by releasing intact, whole mitochondria^11^—possibly contributing to the differing levels of cf-mtDNA and ex-Mito observed between plasma (anticoagulated) and serum (coagulated)^12–16^. In one study, plasma samples processed using filtration and high-speed centrifugation resulted in the release of whole ex-Mito (either free or in large EVs), which made up approximately 99% of all particles found in plasma^13^. However, when platelet activation was avoided, free and EV-encased ex-Mito made up 76-80% of plasma particle types, while the amount of mtDNA detected decreased 67-fold^13^. Furthermore, the cellular sources of ex-Mito in blood are likely diverse, as recent reports demonstrated that at least some parenchymal cells, such as adipocytes, can release their mitochondria into blood. Saliva also contains cf-mtDNA^9,17^, suggesting that non-platelet sources, such as salivary glands or other buccal cells, may contribute to saliva cf-mtDNA abundance^17^. Interestingly, salivary cf-mtDNA has been show to increase several-fold in response to acute mental stress^15^.

These findings suggests that 1) free mitochondria and EV-Mito containing whole mitochondrial genomes may largely account for mtDNA detected across plasma samples, 2) the preparation of plasma samples with platelet activation can lead to the release of whole mitochondria, and 3) there may be diverse ex-Mito in biofluids. These considerations make it important to distinguish between mtDNA fragments and whole mitochondria when assessing cf-mtDNA. They also highlight the need to define the nature of circulating mitochondrial components across human biofluids.

In this study, we collected venous blood and saliva from ten healthy adult volunteers, isolated cf-mtDNA-rich fractions by differential centrifugation and size-exclusion chromatography, and imaged the resulting material using transmission electron microscopy. We provide a catalogue and image bank of cf-mtDNA-enriched particles from six human biofluids.

Consistent with the emerging literature, our findings reveal the presence of structures that morphologically resemble mitochondria in multiple biofluids, with the highest levels seen in plasma followed by saliva. The resulting image inventory expands our knowledge of the wide range of circulating particles in human blood and saliva, and serves as a foundation to inform biofluid selection and technical considerations in future studies.

## 2. Methods

### 2.1 Blood and Saliva Collection

Saliva and venous blood were collected from ten healthy adult volunteers who gave informed consent. The study protocol was approved by New York State Psychiatric Institute Institutional Review Board protocol #7618. Blood was drawn with a 21-gauge needle (BD #367281), and 66 mL of blood was collected into 13 tubes. Participants were not required to have fasted. Blood samples were drawn consecutively in the following order: citrate (2 x 2.7 mL) (BD #363083), heparin (6.0mL) (BD#367878), and EDTA (6.0mL) (BD #366643) for plasma samples, and red top (6.0 mL) (BD #367814) and gold top (6.0mL) (BD #367989) for serum samples. After collection, all tubes were inverted 10-12 times to ensure thorough mixing. Saliva was sampled concurrently using a Salivette cotton swab (Sarstedt #51.1534.500) on the tongue for 5 minutes beginning once the blood started to flow into collection tubes.

### 2.2 Cell-free Plasma, Serum, and Saliva Preparation

Blood samples were processed shortly after collection with an initial centrifugation of 1,000g for 5 minutes (10 minutes for gold top serum tubes). Serum samples were incubated at room temperature for 30 minutes to ensure coagulation prior to initial centrifugation. Both plasma and serum supernatants were transferred to 1.5 mL Eppendorf tubes and centrifuged a second time at 5,000g for 10 minutes at room temperature to pellet and remove the remaining platelets. The resulting plasma/serum was then aliquoted in 150μL aliquots and stored at -80°C for later use. Salivettes were immediately centrifuged at 1,000g for 5 minutes at room temperature; the resulting supernatant was transferred to 1.5mL Eppendorf tubes and stored at -80°C. Prior to use, saliva samples were thawed and centrifuged for a second time at 5,000g for 10 minutes. 30μL of each sample was reserved in a 96-well plate for later qPCR of cf-mtDNA^18^.

### 2.3 Isolation of EVs through Size Exclusion Chromatography (SEC)

EVs from the sixty plasma, serum, and saliva samples were isolated using Izon’s qEV size-exclusion chromatography system (**Figure 1A**). This machine separated particles based on their size as they passed through a column of resin, enabling the delivery of highly purified EVs.

**Figure 1.**
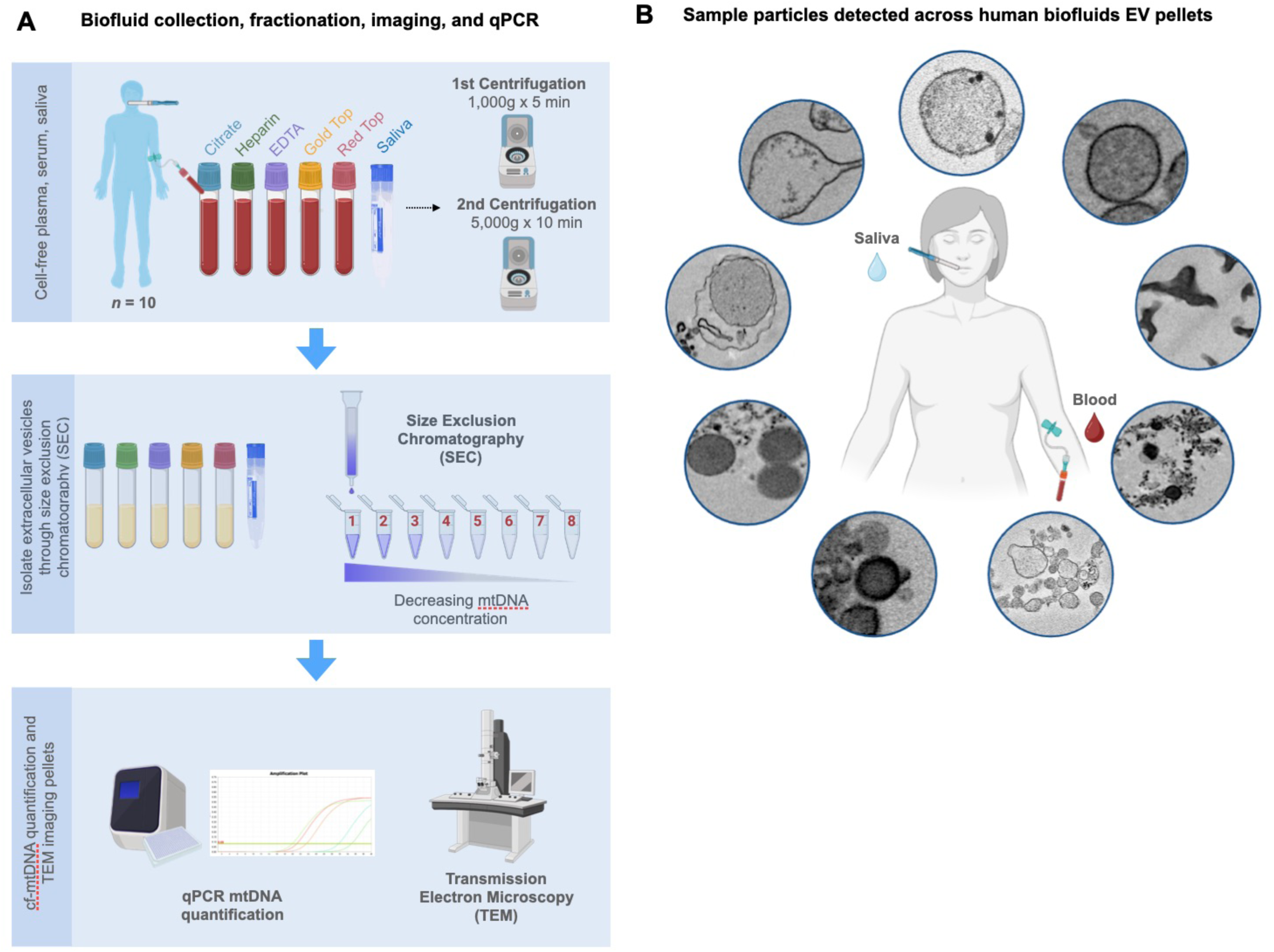
Overview of EV extraction and analysis from plasma, serum, and saliva. **(A)** Experimental protocol starting with blood and saliva collection and cf-plasma, serum, and saliva preparations, followed by purification of material through size exclusion chromatography (SEC). Mitochondrial DNA quantification by lysis (MitoQuicLy) was performed on all biofluid samples, pooled SEC fractions, and supernatant; and isolated pellets were fixed in glutaraldehyde for transmission electron microscopy. For a detailed protocol, see Supplemental Figure 1. **(B)** Schematic of the catalog of circulating particles across human saliva and blood-derived biofluids.

These EVs were distributed in descending size order into eight 1.5mL Eppendorf tubes, with the highest concentration of fractions contained within the first four tubes delivered. The supernatant was aliquoted for later qPCR. The Izon EV fractions were then centrifuged at 18,213g for 30 min at 4°C to form visible pellets, and a formaldehyde fixative was added to stabilize protein structures. These pellets were sent to the Electron Microscopy Core Facility at Weill Cornell Medical Center for transmission electron microscopy analysis.

### 2.4 cf-mtDNA Quantification using qPCR by Lysis

To validate that the centrifuged Izon fractions and pellets sent to the Imaging Core Facility contained highest number of cf-mtDNA copies, we isolated DNA via alkaline lysis and performed qPCR on the reserved 30μL of plasma, serum, saliva, supernatant, and eluate samples. In a 96-well semi-skirted PCR plate, 10μL of each sample was added to 190μL lysis buffer (6% Tween-20 [Sigma #P1379], 114 mM Tris-HCl pH 8.5 [Sigma #T3253], and 200μg/mL Proteinase K [ThermoFisher #AM2548]). After sealing and mixing the plate, the sample/lysis buffer mixture was then incubated overnight at 55°C for 16h in a standard thermocycler, followed by Proteinase K inactivation step at 95°C for 10 minutes, and held at 4°C until qPCR.

Cf-mtDNA concentration was then quantified by qPCR. The ND1 amplicon targets the mtND1 Gene (ENSG00000198888.2) in the mitochondrial genome^18^. A master mix was prepared for each experimental plate by combining 2xTaqMan Universal Mastermix Fast (Life Technologies #4444965) with 10μM ND1 Primers F+R, 10μM B2M Primers F+R, 5μM ND1 Probes-VIC, and 5μM B2M Probes-FAM (idtDNA.com)^18^. The final concentrations of primers were 300nM, and the final concentrations of probes were 100nM. 12μL of master mix and 8μL of the lysate (input mtDNA) were dispensed into each well of the 384-well qPCR reaction plate (ThermoFisher # 4309849)^18^<u>10/15/25 1:46:00 PM</u>. All reactions were carried out in triplicate.

The plates were sealed and pulse-spun to 1,000g, and qPCR was performed on the QuantStudio 7 Flex Real-Time PCR System (ThermoFisher #4485701)^18^.

### 2.5 Statistical and Image Analysis

Averages, standard deviations, and coefficients of variation (CV) were computed between qPCR triplicates using Microsoft Excel. Outlier wells that resulted in CVs greater than 10% between triplicates were discarded. Statistical analyses of qPCR results were performed using GraphPad Prism (version 9). Electron microscopy images of the extracellular vesicles in each sample were analyzed using FIJI analysis software through image segmentation, cell counter, and particle analysis tools.

## 3. Results

### General appearance of pelletable structures from human biofluids

Imaging of EV pellets from human blood and saliva showed a heterogeneous population of particles, including electron-dense, membrane-bound structures consistent with hallmarks of mitochondrial ultrastructure (**Figure 1B**). Aiming to create an inclusive catalog of circulating vesicle types and other structures, we identified 14 distinct particle types across citrate-, heparin-, and EDTA-plasma; red top and gold top serum; and saliva samples (**Figure 2**). The overall appearance of biofluid pellets differed considerably across human biofluids (**Figure 3**), including in the overall size, diversity, and electron density of structures. **Figure 4** highlights the appearance of mitochondria-like structures, along with a wide-range of vesicles, across biofluids from two participants. Eukaryotic human cells contain only two types of double membrane-bound structures: the nucleus and mitochondria. Because some of the double membrane-bound structures contained internal cristae-like membranes, the type “F” double-membrane structures illustrated in **Figure 4**, which were observed in some but not all human biofluids, are likely circulating ex-Mito.

**Figure 2.**
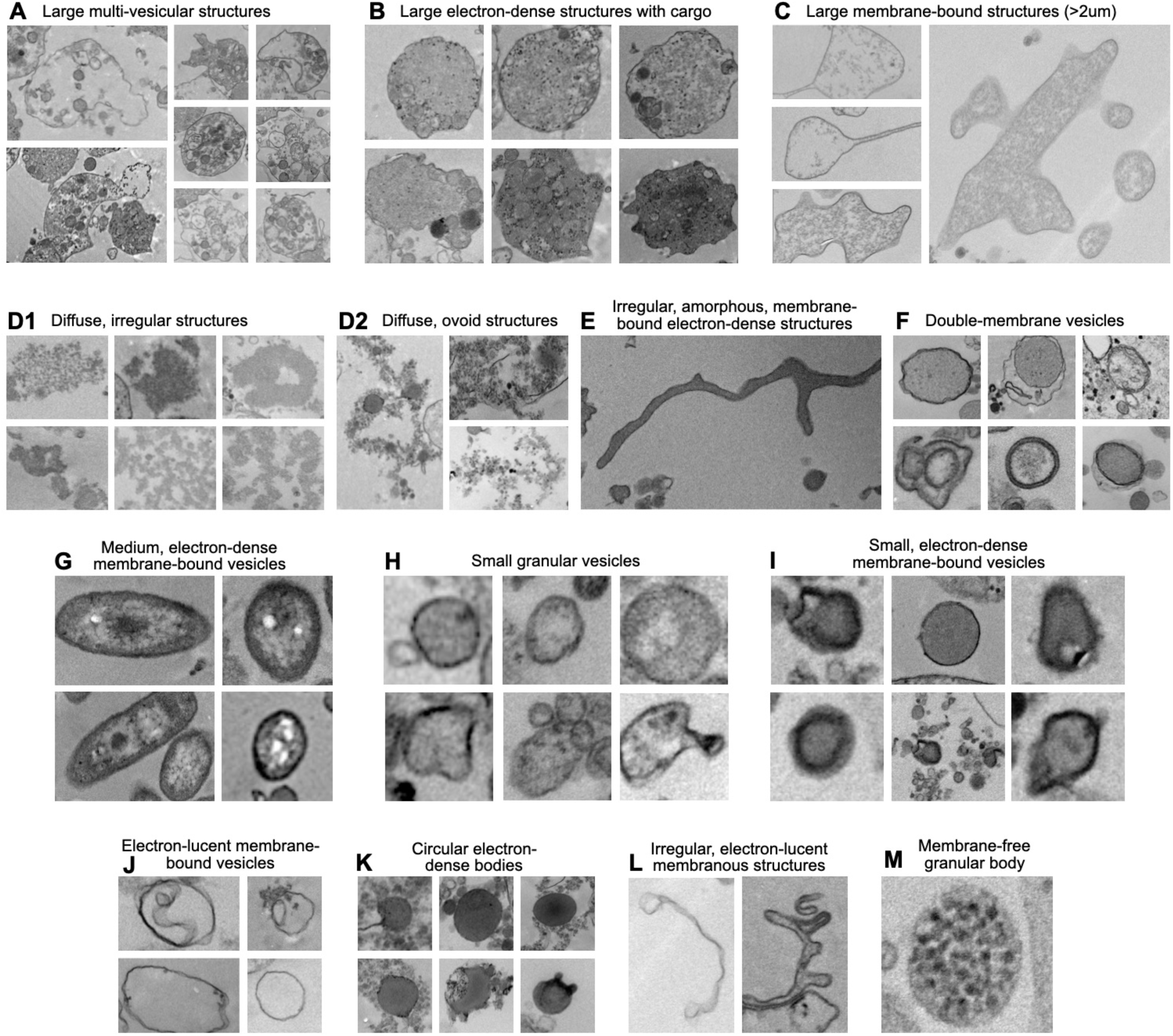
Categorization of particle types in human plasma, serum, and saliva. Complete inventory of distinct particle types identified in 109 Transmission Electron Microscope (TEM) images across biofluids. Each letter corresponds to a particle type that was determined by grouping particles with similar morphology and electron density. Identified particle types include: **(A)** Large (∼2.5µm) membrane-bound, multi-vesicular structures; **(B)** Large (∼2.2µm) electron-dense structures, with occasional vesicular cargo; **(C)** Large (∼2.2µm) membrane-bound, electron-lucent but granular structures; **(D1)** Diffuse, irregular structures; **(D2)** Diffuse, ovoid structures; **(E)** Irregular, amorphous, membrane-bound electron-dense structures; **(F)** Double-membrane vesicles; **(G)** Medium (∼0.3µm) membrane-bound electron-dense vesicles complex ovoid structures with compartmentalized granular material, and electron dense external membrane; **(H)** Small (∼0.15µm) granular vesicles; **(I)** Small (∼0.1µm) membrane-bound electron-dense vesicles; **(J)** Small (∼0.1µm) membrane-bound electron-lucent vesicles; **(K)** Small (∼0.1µm) circular, electron-dense amorphous bodies; **(L)** Small (∼0.07µm) membranous irregular structures; **(M)** Small (∼0.06µm) circular membrane-free granular body.

**Figure 3.**
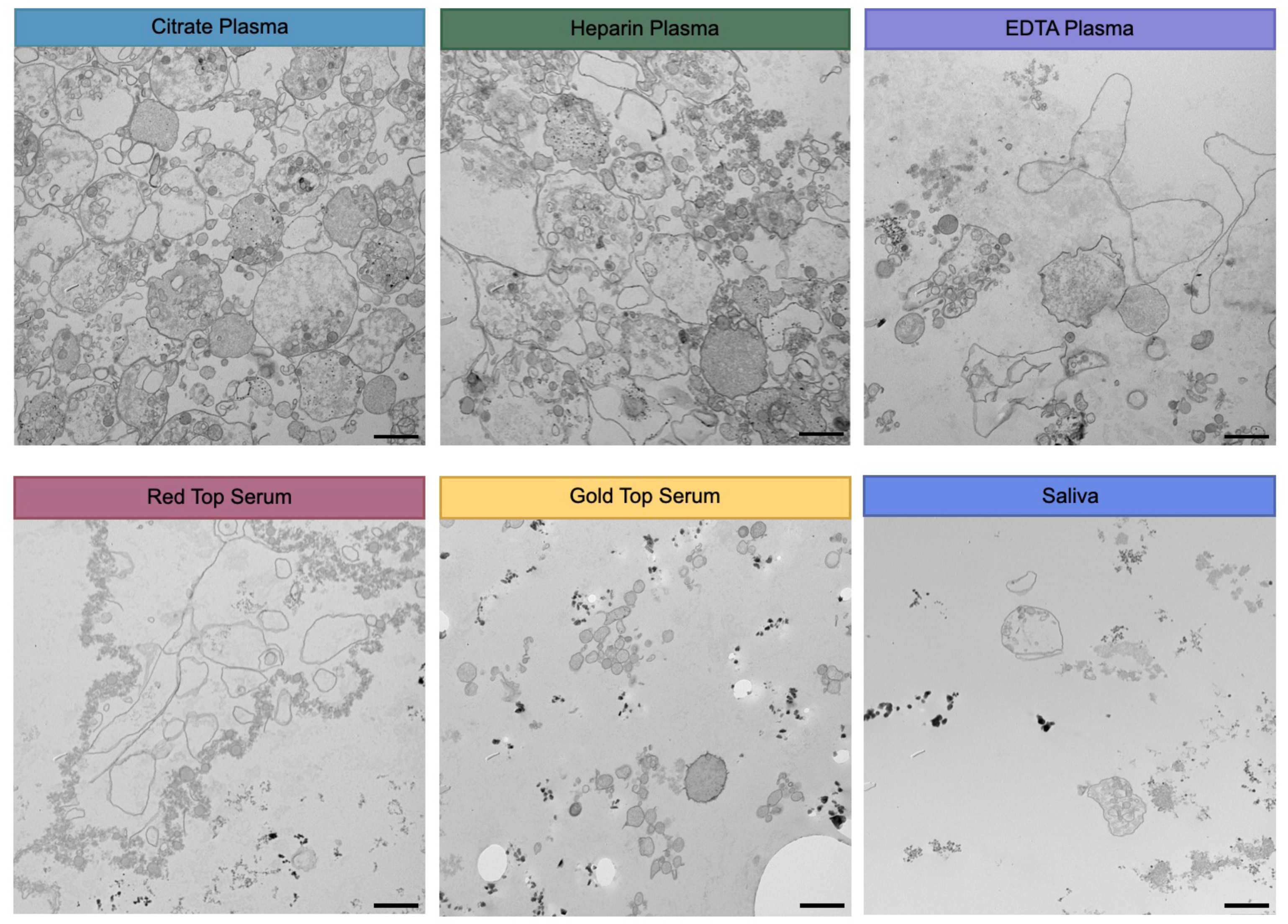
Electron microscopy of extracellular vesicles in plasma, serum, and saliva. 15,000x magnification transmission electron microscopy images from representative study participants; citrate, heparin, and EDTA plasma, red top and gold top serum, and saliva. Images were chosen to provide a representative overview of the extracellular environment of each biofluid. Scale bar, 1um.

**Figure 4.**
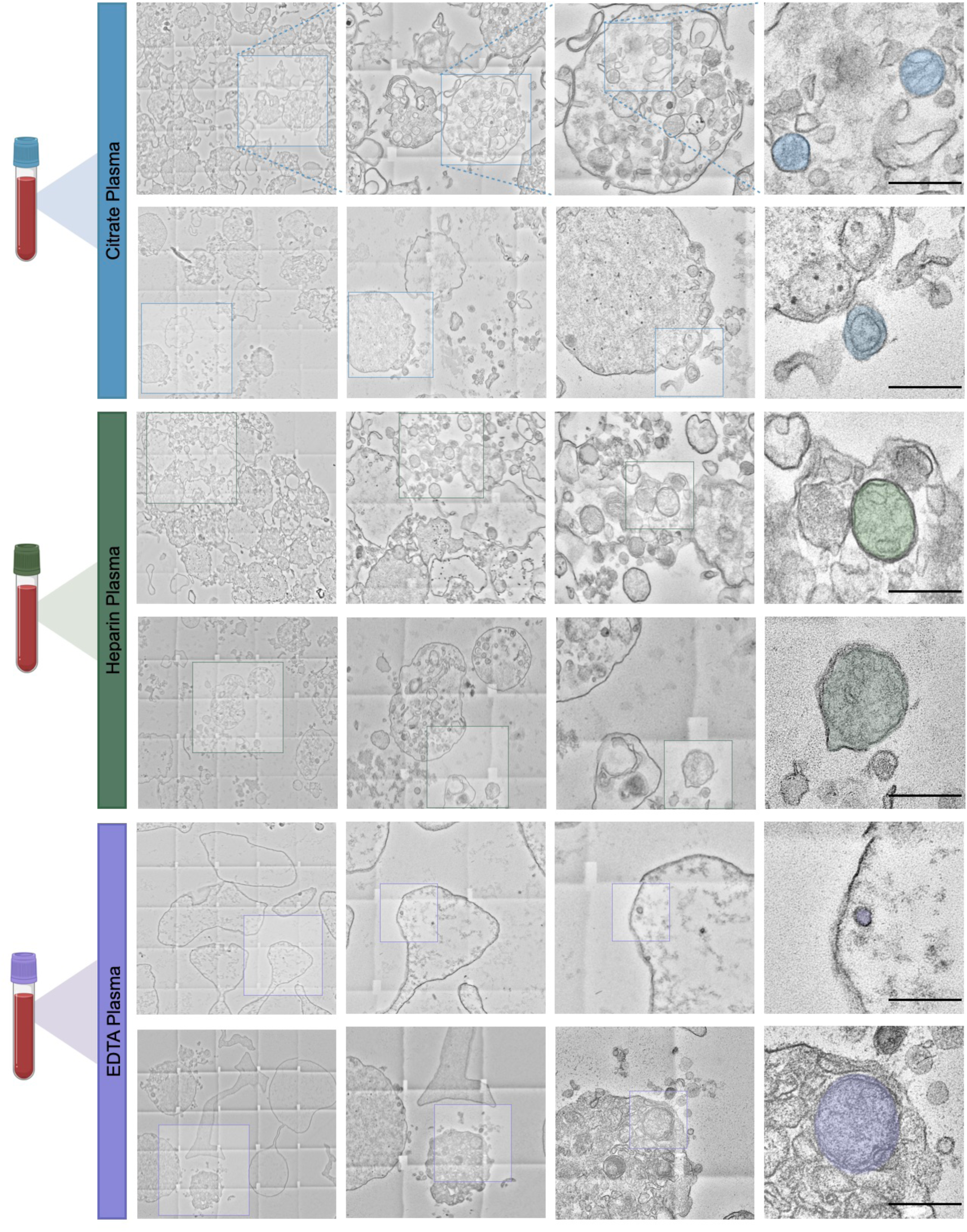
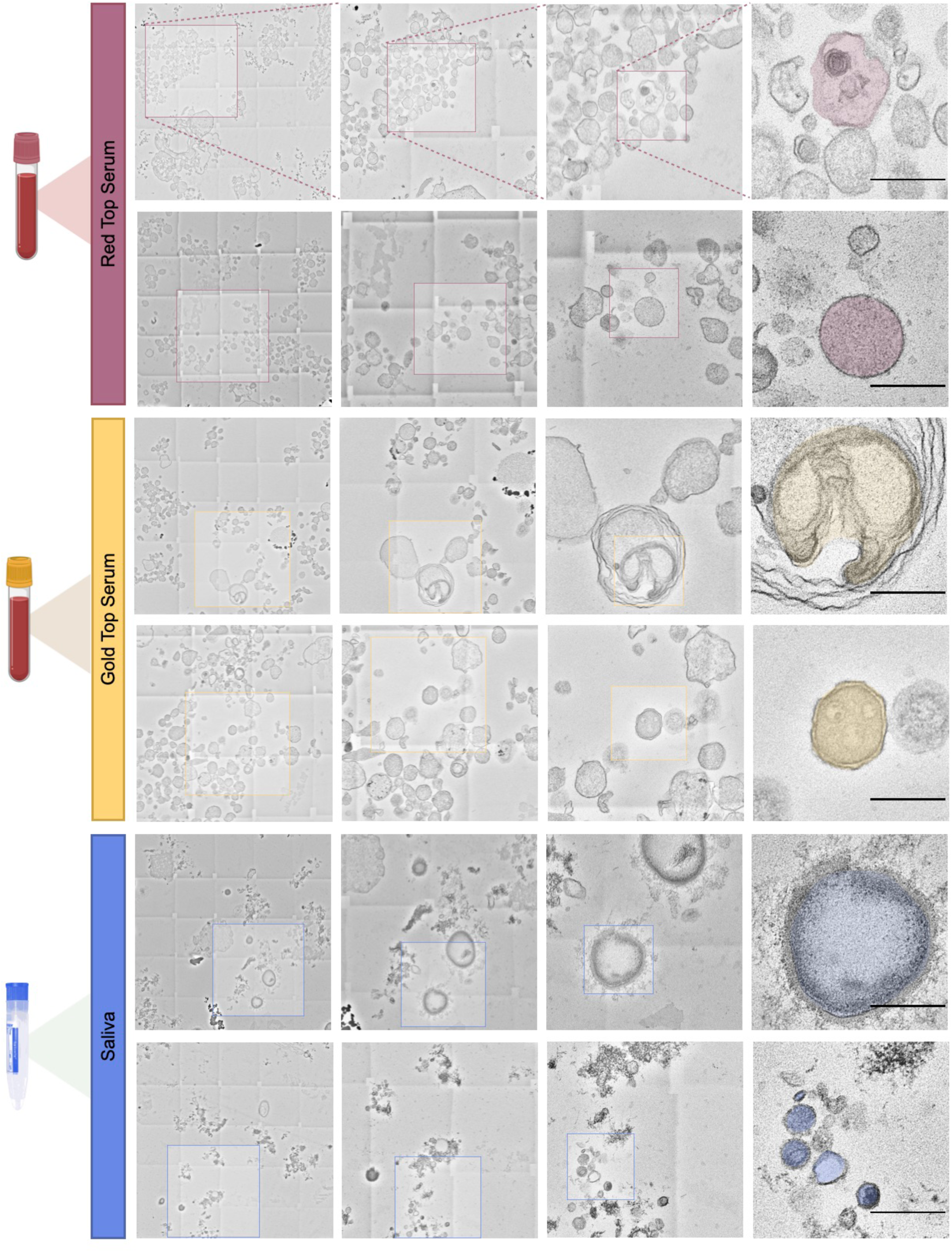
Electron microscopy magnification of extracellular vesicles in plasma, serum, and saliva. 15,000x magnification transmission electron microscopy images from representative study participants; citrate, heparin, and EDTA plasma, red top and gold top serum, and saliva. Images were selected to provide an overview of diverse particle types found throughout human biofluids. A more detailed categorization of each particle type can be found in Figure 4. Scale bar, 5um.

### Size and electron density of circulating structures

We systematically analyzed the size and electron density for each of the 14 distinct particle types (**Figure 5**). Large (∼2.2-2.5μm) membrane-bound, multi-vesicular structures (Types A and B, respectively) were the largest observed, ranging from ∼0.6 to 7μm (**Figure 5A**). Most of these large structures were consistent in morphology with platelets and contained several vesicular structures as cargo. The smallest particles were small membrane-bound electron-dense vesicles (Type I), all being less than 0.1μm.

**Figure 5.**
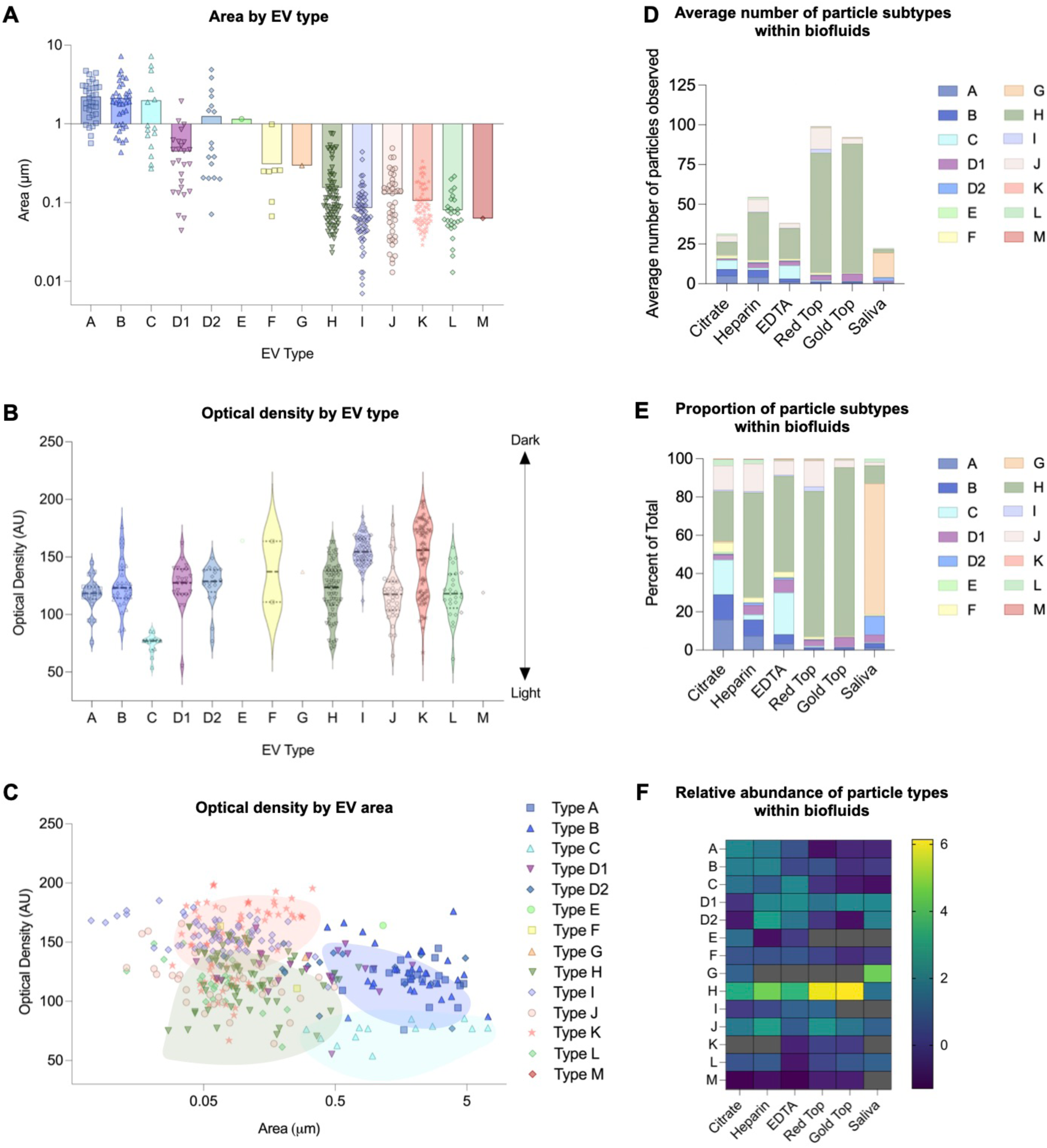
Morphological, optical, and quantitative characterization of extracellular vesicles in plasma, serum, and saliva. **(A)** Column scatter plot of distinctions in area of each particle type with logarithmic area scale. **(B)** Violin plot of optical densities of each particle type with standard optical density scale. On this scale, 0 represents white, and 250 represents black. **(C)** Scatter plot of area vs optical density of each particle category with logarithmic area scale. Each shape/color corresponds to a particle type as seen in the key on the right-hand side. **(D)** Average number of particle subtypes observed in each biofluid. Each color corresponds to a particle type as denoted in the key on the right-hand side. **(E)** Proportion of particle subtypes within each biofluid. Each color corresponds to a particle type as labeled in the right-hand key. **(F)** Heatmap of the relative abundance of particle types within each biofluid; each letter corresponds to a particle type shown in Figure 2.

The darkest particles were small (∼0.1μm) membrane-bound, electron-dense vesicles, and circular, electron dense amorphous bodies (Types I and K, respectively) (**Figure 5B**).

Conversely, the least dense structures were large (∼2.2µm) membrane-bound, electron-lucent but granular structures (Type C). By analyzing optical density relative to EV area, we found a modest trend showing that larger EVs (≥0.5µm) tended to exhibit lower optical density compared to smaller EVs (≤0.5µm) (r=-0.27, p<0.0001; **Figure 5C**).

While structural properties provide insight into EV composition, understanding their distribution across different biofluids is critical to assessing their biological relevance. To address this, we analyzed the relative abundance of each particle type across biofluids (**Figure 5D**). Small (∼0.15µm) granular vesicles (Type H) were the most common particle type across both anticoagulated plasma and coagulated serum biofluids, but not in saliva. In plasma, Type H particles accounted for more than 50% of all particles observed in heparin- and EDTA-tubes. In saliva, the majority (67%) of particles were small (∼0.1µm) circular, electron-dense amorphous bodies (Type G).

ex-Mito-like, double-membrane Type F vesicles were present in all biofluids. They represented on average 10.28% of particle subtypes in plasma (approximate total particle count = 1,122) , compared with approximately 1.71% in serum (approximate total particle count = 1,723) , and 0.89% in saliva (approximate total particle count = 107) (**Figure 5E**).

Having identified and analyzed these distinct particle types in aggregate, to determine whether they were linked to mitochondrial content we then examined associations between their abundance and cf-mtDNA levels measured in each participant. We performed size-exclusion chromatography (SEC) and quantified cf-mtDNA in each fraction, across all biofluids, as well as in the total (pre-column) biofluid (**Figure 6A**). mtDNA content was significantly higher in early SEC fractions (containing the largest particles) of citrate and heparin plasma, and nearly absent from post-centrifugation supernatant (**Figure 6A**, *far right column*), further confirming the membrane-bound nature of cf-mtDNA.

**Figure 6.**
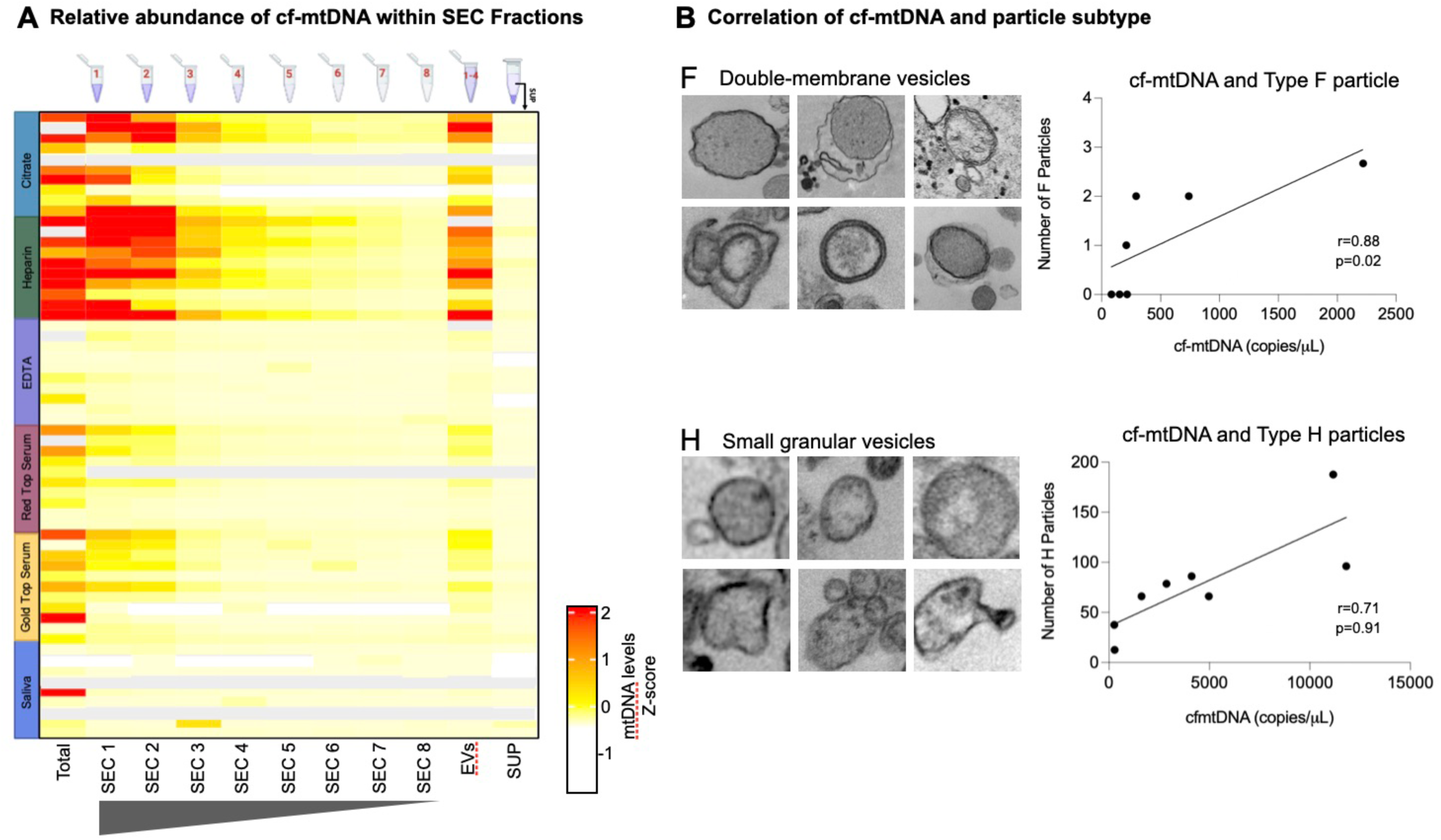
Quantification of cf-mtDNA within samples. **(A)** Heatmap of the relative abundance of cf-mtDNA levels in each SEC fraction of each biofluid. Grey cells indicates missing samples. **(B)** Spearman’s correlation of cf-mtDNA copies/µL and number of particles in F and H particle types, which showed the largest effect sizes. Spearman rs, unadjusted P values.

Although critically underpowered for this kind of quantitative analysis, we correlated the amount of cf-mtDNA in each person (n=7-10) and biofluid (n=6) with all particle types. These preliminary analyses showed that both ex-Mito-like Type F particles in EDTA plasma (r=0.75, p=ns; **Figure 6B**, *top*) and small (∼0.15μm) granular Type H vesicles in red top serum (r=0.81, p<0.05; **Figure 6B**, *bottom*) were most likely to be associated with cf-mtDNA levels. These correlations suggest that approximately 55-65% of the variation in cf-mtDNA levels is explained by variation in Type F and H vesicles. As a potentially notable observation, of the 7 individuals with both cf-mtDNA and electron micrographs available, the three individuals in whom we found no Type F particles had the lowest EDTA plasma cf-mtDNA.

## 4. Discussion

Taken together, these findings combining imaging and cf-mtDNA quantification are consistent with the emerging notion that whole mitochondria released from one or more cellular sources exist and are detectable, albeit in different forms, across different biofluids. These results emphasize the importance of blood tube type selection for examining ex-Mito biology in humans.

Some studies, including from our group, previously assumed that circulating mtDNA existed as cell-free and membrane-free mtDNA, interpreting associations between cf-mtDNA levels and inflammatory disease states as evidence of its bacteria-like, pro-inflammatory effects^19–23^. Building on that logic, cf-mtDNA initially emerged as a biomarker for diagnostic applications across various pathologies, including depression, cancer, diabetes, Systemic Lupus Erythematosus (SLE), and SARS-CoV-2 infection^5,24–30^. These findings have contributed to a widely held notion that cf-mtDNA is pro-inflammatory^31–39^.

However, this may not be the case if cf-mtDNA is packaged within whole mitochondria or EVs^13^. For cf-mtDNA to be inflammatory, it must be accessible to pro-inflammatory receptors that activate inflammatory responses, such as TLR9^9,26^. However, if cf-mtDNA is contained in vesicles or whole mitochondria, it would only become accessible to TLR9 following endocytosis and other complex intracellular processing that exposes free mtDNA to the cytosol^9^.

Interestingly, a series of recent studies indicate that directly injecting isolated mitochondria by intraperitoneal or intravenous routes is either not associated with inflammation or has anti-inflammatory effects^40–44^, challenging the idea that cf-mtDNA is exclusively pro-inflammatory. Further, evidence suggests that cf-mtDNA is present in the blood even when there is no systemic inflammation^9^.

It is estimated that there are between 200,000 and 3.7 million intact, cell-free mitochondria circulating per mL of plasma, potentially accounting for over 90% of cf-mtDNA in healthy women and men^9^. This suggests that circulating mtDNA has a biological function beyond serving as an inflammatory marker, though the functional effects of cf-mtDNA may be disease and/or context-specific. One of these functional effects includes interorgan mitochondria transfer^3,45^. ex-Mito have been shown to circulate in human biofluids post-surgery (e.g., in cerebrospinal fluid following subarachnoid hemorrhage), suggesting an alternative role in healing and repair^47^. Beyond vesicles, multiple cells-of-origin may contribute to ex-Mito in blood, including platelets and adipocytes^3,4,11^. An interesting future direction would be to determine the diversity of cellular origins for cf-mtDNA and their relative distributions within and outside of circulating ex-Mito.

This catalogue of cf-mtDNA-enriched circulating structures from six human biofluids serves as a resource to support the design of future research on mitochondria transfer, cf-mtDNA, and ex-Mito biology.

## Author contributions

J.M. and M.P. designed and conceptualized the study. J.M. and S.G.F. processed samples and analyzed data. S.G.F. analyzed images, performed the quantitative analysis, and drafted the figures. S.V. finalized data analysis, visualization and figures. S.V., S.G.F., and M.P. drafted the manuscript. J.R.B. and S.R. edited and provided feedback on the manuscript. M.P. supervised the project. All authors provided feedback on the manuscript.

## Acknowledgements

This work was supported by NIH grant R01MH119336, the Wharton Fund, and Baszucki Group to M.P.

## Data Availability

All images and data are available upon request. All images can be accessed at https://figshare.com/s/af2348b12c5950f87ae8.

---

**Supplemental Figure S1.**
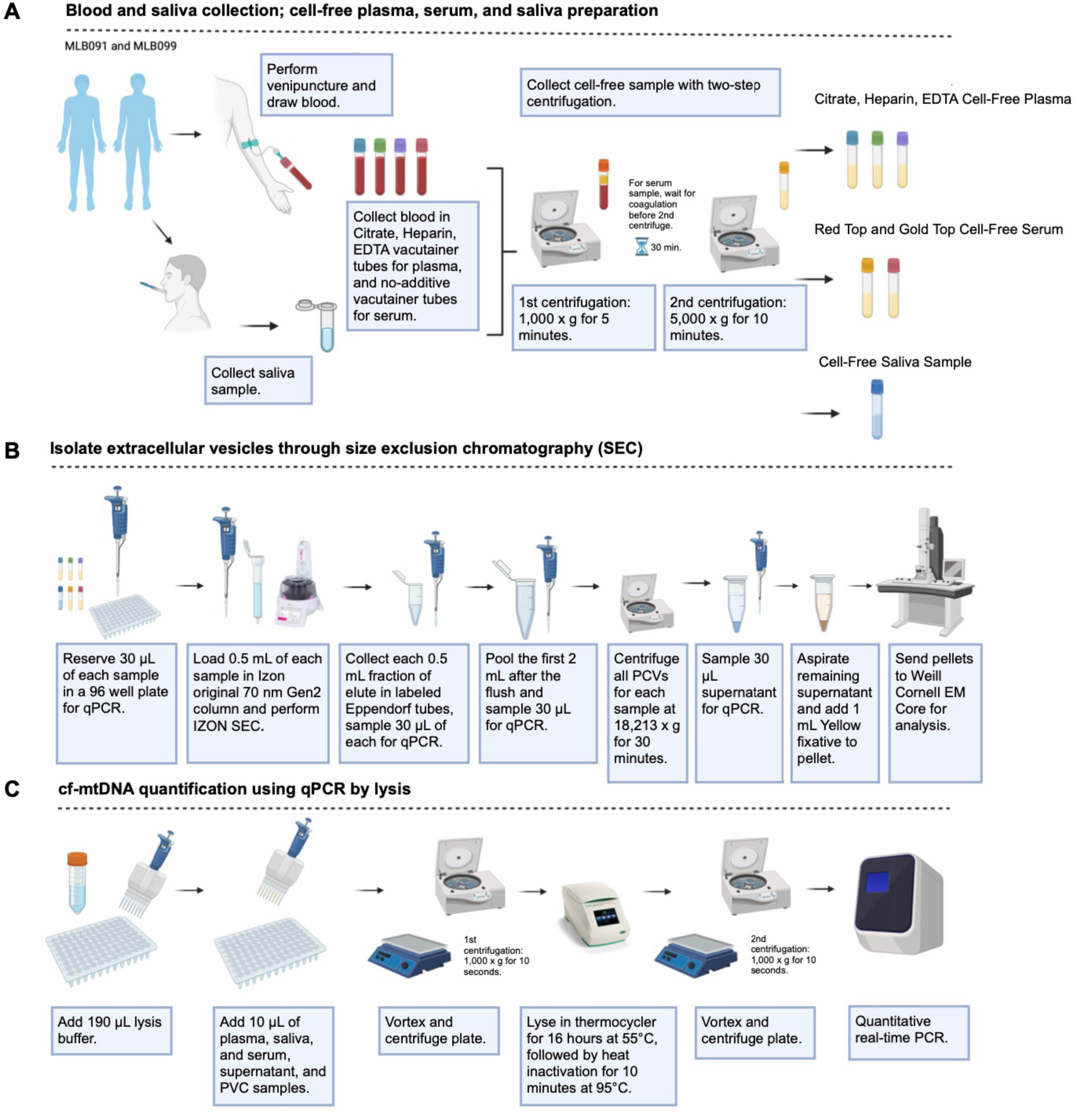
Detailed schematic of experimental protocol. **(A)** Blood and saliva collection and cf-plasma, serum, and saliva preparation. **(B)** Isolation of extracellular vesicles through size exclusion chromatography using Izon’s SEC protocol, with the isolated pellets fixed and sent to Weill Cornell EM core for analysis. **(C)** To validate that the pellets sent to Weill had the highest possible amounts of cf-mtDNA, qPCR by lysis was performed of all biofluid samples, pooled SEC fractions, and supernatant.

